# Use of Chinese herbs to treat symptoms likely related to COVID-19: Survey analysis of licensed acupuncturists in the United States

**DOI:** 10.1101/2022.06.23.22276829

**Authors:** Belinda J. Anderson, Melissa Zappa, Brent D. Leininger, Lisa Taylor-Swanson

**Affiliations:** Pace University, College of Health Professions, 163 Williams St, New York, NY 10038, USA; Albert Einstein College of Medicine, 1300 Morris Park Ave, Bronx, NY 10461, USA; Huntsman Cancer Institute, University of Utah, Salt Lake City, Utah, USA; Integrative Health & Wellbeing Research Program, Earl E. Bakken Center for Spirituality & Healing, University of Minnesota, 420 Deleware St SE, Minneapolis, MN 55455; College of Nursing, University of Utah, 10 S 2000 E, Salt Lake City, Utah, USA

**Keywords:** COVID-19, Chinese herbal medicine, complementary medicine, pandemic

## Abstract

**Objective:** The objective of this study was to examine the prescribing of Chinese herbal medicine (CHM) by licensed acupuncturists in the United States during the COVID-19 pandemic.

**Methods:** A 28-question survey with nine branching questions was disseminated through collegial networks, paid advertisements, and a study website in April-July 2021. Participants indicated they were licensed acupuncturists that treated more than five patients for symptoms likely related to COVID-19 to gain entry to the full survey. Surveys were undertaken electronically through the Research Electronic Data Capture (REDCap) system.

**Results:** The survey was undertaken by 103 participants representing all US geographic regions, and had an average of 17 years in practice. Sixty-five percent received or intended to receive the COVID-19 vaccine. Phone and videoconference were the predominant methods of patient contact; granules and pill forms of CHM were the most commonly prescribed. A wide variety of information sources were used in devising patient treatments inclusive of anecdotal, observational, and scientific sources. Most patients were not receiving biomedical treatment. Ninety-seven percent of participants reported that they had no patients die of COVID-19, and the majority reported less than 25% of their patients developed long hauler syndrome (post-acute sequelae SARS-CoV-2 infection).

**Conclusions:** This study demonstrates that licensed acupuncturists were treating COVID-19 infected individuals in the US during the early stages of the pandemic, and for many such patients this was the only therapeutic intervention they had access to from a licensed healthcare provider. Information disseminated from China through collegial networks, along with published sources including scientific studies, informed the approach to treatment for the vast majority of the acupuncturists surveyed. This study provides insight into an unusual circumstance in which clinicians needed to establish evidence-based approaches to the treatment of a new disease in the midst of a public health emergency.

**Strengths and Limitations of the Study:**

- Detailed survey that was widely disseminated through paid advertisements and a study web site
- Data was only collected from licensed clinicians who had treated more than five patients with symptoms likely related to COVID-19
- Survey was undertaken through the Research Electronic Data Capture (REDCap) system
- Statistical analysis was undertaken to determine associations between question responses
- The survey was not psychometrically tested

## Introduction

One of the most concerning aspects of coronavirus disease (COVID-19) during the first year of the pandemic was that there were no biomedical antiviral treatments. In China use of traditional Chinese medicine (TCM) for COVID-19 was undertaken both alone and in combination with biomedical symptomatic treatments,^1,2^ and the Chinese government established a prevention and control strategy based on TCM and biomedicine.^3^ Chinese herbal medicine (CHM) was an important part of this strategy because it had been used extensively for numerous prior epidemics, which also significantly shaped theories and treatment approaches throughout the history of the use of CHM.^2,4,5,6,7^. More recently CHM was successfully utilized during the 2003 SARs epidemic and the 2009-10 flu season associated with H1N1 virus.^8,6,9^ The broad symptom picture that COVID-19 infected patients can present with are encompassed by TCM diagnostic patterns and organ relationships,^1^ and the use of TCM theories has provided insight into the pathology associated with COVID-19 infection.^10^

East Asian medicine (EAM) has been practiced throughout Asia for thousands of years, and more recently all over the world. EAM encompasses several different modalities including but not limited to acupuncture, CHM, moxibustion, cupping, and Tui Na (Chinese medical massage). A very large body of clinical research supports the efficacy and effectiveness of acupuncture,^11^ which is now in widespread use throughout the US and covered by many private insurers and Medicare for lower back pain.^12,13^ A smaller body of research evidence supports the efficacy and effectiveness of the other EAM modalities, including CHM. Development of the knowledge base supporting CHM was largely undertaken in China throughout the last several millennia, and represents both classical and more modern approaches to the therapeutic use of the various constituents in CHM pharmacopeia.^14^ The modern approach to EAM in China is referred to as Traditional Chinese medicine (TCM) and is distinguished from the preceding approaches commonly referred to as classical Chinese medicine.^15^

The CHM pharmacopeia consists of over 500 different constituents, which are largely of plant origin, along with others of animal and mineral origin. The widespread acceptance of CHM was significantly increased following the Nobel Prize award to You-you Tu for the discovery of artemisinin, a therapeutic drug for malaria derived from the plant *Artenisia annua*, which is a constituent of the CHM pharmacopeia. It is estimated that more than one-third of clinical drugs are extracted and/or derived from CHM resources.^16^

CHM prescriptions usually consist of 3-20 different constituents, which are called herbal formulas.^17,18^ Practitioners often start with a commonly used formula from documented sources (classical or modern) and then modify by adding and/or removing specific constituents such that the final formula is specific for the presenting condition of individual patients. Choice of formulas and individual constituents is made within the context of East Asian medical diagnostic and treatment theory, along with an understanding of the actions and effects of formulas and constituents. CHM can be taken internally as a decoction (CHM is cooked in water and strained), or a powder (often termed granules) that is dissolved in water, or in pill form.

Given the use and applicability of TCM for managing the COVID-19 pandemic in China, information about its use, especially the use of CHM, spread rapidly to other countries in the early stages of the pandemic. In the United States (US) licensed TCM practitioners (usually referred to as licensed acupuncturists) accessed this information through collegial networks, and through information distributed through continuing education providers, CHM companies, and East Asian medicine journals. One significant source of such information was through the Lotus Institute for Integrative medicine^19^ who also disseminated information from Dr. John Chen, an internationally recognized CHM expert who has authored many textbooks on this topic.^20,21,18^ In the US, state laws vary as to regulations associated with licensed acupuncturists prescribing CHM. Some states require licensed acupuncturists to pass a certification exam provided by the National Certification Commission for Acupuncture and Oriental Medicine^22^ that is specific to CHM. Other states have no regulations associated with prescribing and/or recommending CHM.

By mid-2020 publications started to appear in scientific journals describing observational studies and randomized controlled clinical trials assessing the effectiveness and efficacy of CHM for COVID-19 symptoms. In a systematic review and meta-analysis published in July 2020^23^ Xiong and colleagues pooled the data from 18 trials involving 2275 patients and concluded that there was suggestive evidence that CHM may be beneficial for the treatment of COVID-19 through improvements in clinical symptoms, imaging outcomes, laboratory indicators, shortening the course of the disease, and reducing the number of severe cases. Since that time many more randomized controlled trials of CHM for COVID-19 symptoms have been published,^9,2^ further suggesting CHM may be beneficial for treating COVID-19 symptoms.

In many states throughout the US, licensed acupuncturists were not considered essential workers during the early stages of the COVID-19 pandemic and were required to close their offices due to lockdown mandates. Many continued to provide medical services to their patients through telehealth^24^ and CHM medicine was one of the main therapeutic interventions that were offered. The importance of their role in providing treatment was accentuated by the lack of effective biomedical treatments and the rapid rise in infections and deaths. Given that this was a new disease, practice guidelines for using CHM for COVID-19 symptoms were nonexistent at the beginning of the pandemic. This obviously created a significant limitation to an evidence-based approach to treatment. Yet, classical and modern evidence on the use of CHM for infectious disease existed, which informed clinical decision making. Our study was designed to investigate how licensed acupuncturists who were treating patients with symptoms likely related to COVID-19 with CHM made treatment decisions, and how the pandemic impacted their practices.

## Methods

### Survey instrument

We used a cross-sectional study design with the anonymous survey distributed in the Spring/Summer of 2021. The survey questions were devised by the authors through an iterative process of survey draft editing conducted through email and study team meetings. The final survey instrument consisted of 28 primary questions. Nine of these questions had additional branching questions that depended upon the response to the primary question. The survey was not psychometrically tested.

Initial survey questions collected general demographic information. The first two questions were – 1. Are you a licensed acupuncturist in the USA, and 2. Have you treated more than five patients with symptoms that may be related to COVID-19 with Chinese herbal medicine? If the respondents answered no to either of these questions they were discontinued from the survey.

Therefore, the survey only permitted completion by respondents who were licensed acupuncturists in the US, and who had treated more than five patients with symptoms that may be related to COVID-19 with CHM. The following questions after the demographic questions asked about personal infection and vaccine participation, numbers and types of patients treated with symptoms likely related to COVID-19, how patient appointments were conducted, form of CHM prescribed, sources of information used in devising CHM formulas, patients’ use of biomedical treatments, duration of CHM treatment, outcome assessment, side effects, difficulty obtaining herbs, and incidence of long-covid and death among patients.

### Participant recruitment

Survey participants were recruited by disseminating survey invitations among the author’s colleagues, paid advertisements through Acupuncture Today, and by creating a website – www.teamcovidstudy.com. No compensation was provided to participants. At the end of the survey, there was an invitation to participate in an individual interview as part of the qualitative component of this study that is reported in a separate publication.

### Survey implementation

The use and implementation of the survey was approved by the Institutional Review Board of Albert Einstein College of Medicine (Einstein). Information about the survey and informed consent was presented at the beginning of the survey, and survey participation indicated informed consent. The survey was conducted between April 1^st^ 2021 and July 20^th^ 2021. Surveys were undertaken online through the Einstein Research Electronic Data Capture (REDCap) system.

### Statistical analysis

We used descriptive statistics to report the number and percentage of responses to individual survey questions. We used Chi-square or Fisher exact tests to assess potential associations between formal research or evidence-based medicine training and 1) influence by scientific studies; 2) use of biomedical journals to inform COVID-19 herb formulations; and 3) use of outcome instruments. Fisher exact tests were used when expected frequencies of individual cells were less than 5.^25^ We used a two-sample t-test or ANOVA (when more than 2 groups were present) to assess for differences in years of clinical experience based on self-reported 1) use of clinical experience in COVID-19 herb formulation and 2) modifications to COVID-19 herb formulations. All analyses were conducted using Stata 13.1.^26^

## Results

The survey was initiated by 125 respondents. Of the 125, two were not licensed acupuncturists in the US, and 20 had not treated more than five patients with symptoms likely related to COVID-19 with CHM. Of the remaining 103 respondents, 25 partially completed the survey, and 78 completed the survey.

Table 1 presents the demographic data for participating acupuncturists. Fifty-eight percent of the participants had current certification through the National Certification Commission for Acupuncture and Oriental Medicine (NCCAOM), and 8% had had NCCAOM certification previously. Practitioners were representative of all four US geographic regions as defined by the US Census.^27^ Greater representation was from the northeast and west regions. Participants had been in practice for an average of 17 years, with a range of 1 to 40 years. Sixty-eight percent had had formal research training, which had been completed on average 12 years prior (range 0-38 years).

**Table 1.** Demographic data

| NCCAOM certification | n (%) |
| --- | --- |
| Yes | 60 (58.3) |
| No | 35 (34.0) |
| In past | 8 (7.7) |
| Location of practice | n (%) |
| Northeast | 32 (31.4) |
| Midwest | 14 (13.7) |
| South | 15 (14.7) |
| West | 41 (40.2) |
| Years in practice | Mean (sd)<br>[min – max] |
|  | 16.9 (10.3)<br>[1 – 40] |
| Research training | n (%) |
| No | 25 (32.0) |
| Yes | 53 (68.0) |
| Years since completion of training | Mean (sd)<br>[min – max] |
|  | 12.1 (9.9)<br>[0 – 38] |

As shown in Table 2, 17% of the participants had been infected by COVID-19, and 17% had possibly been infected. Of the infected and possibly infected participants, 97% took CHM for their symptoms. Sixty-five percent received or intended to receive the COVID-19 vaccine, and 7% were unsure.

**Table 2.** Practitioner personal COVID-19 experience

| COVID-19 infection of practitioner | n (%) |
| --- | --- |
| Yes | 17 (16.8) |
| No | 67 (66.3) |
| Unsure | 17 (16.8) |
| Use of Chinese herbs for infection/possible infection | n (%) |
| Yes | 33 (97.1) |
| No | 1 (2.9) |
| Received or intend to receive the COVID-19 vaccine | n (%) |
| Yes | 67 (65.1) |
| No | 29 (28.2) |
| Unsure | 7 (6.8) |

The majority of the participants had treated 30 or fewer patients for possible COVID-19 symptoms (Table 3). Participants varied considerably as to the proportion of patients that were treated during the acute initial stage of infection with 13% reporting that all the patients they treated were in this stage, and 15% reporting that less than 5% of their patients were in this stage. The majority of patients that participants treated had already tested positive for COVID-19. Of the participants’ patients that had access to COVID-19 testing, most tested positive.

**Table 3.**
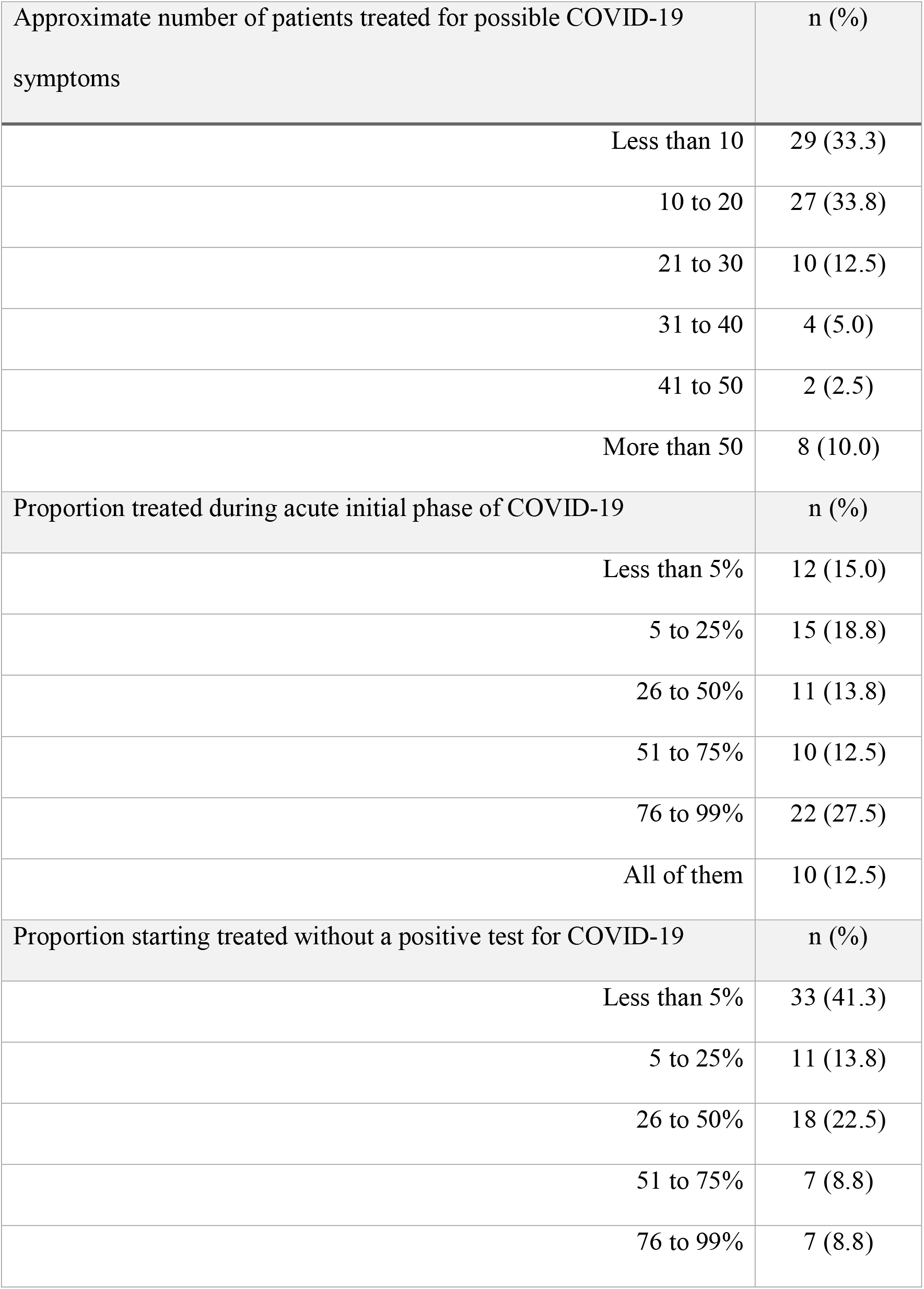
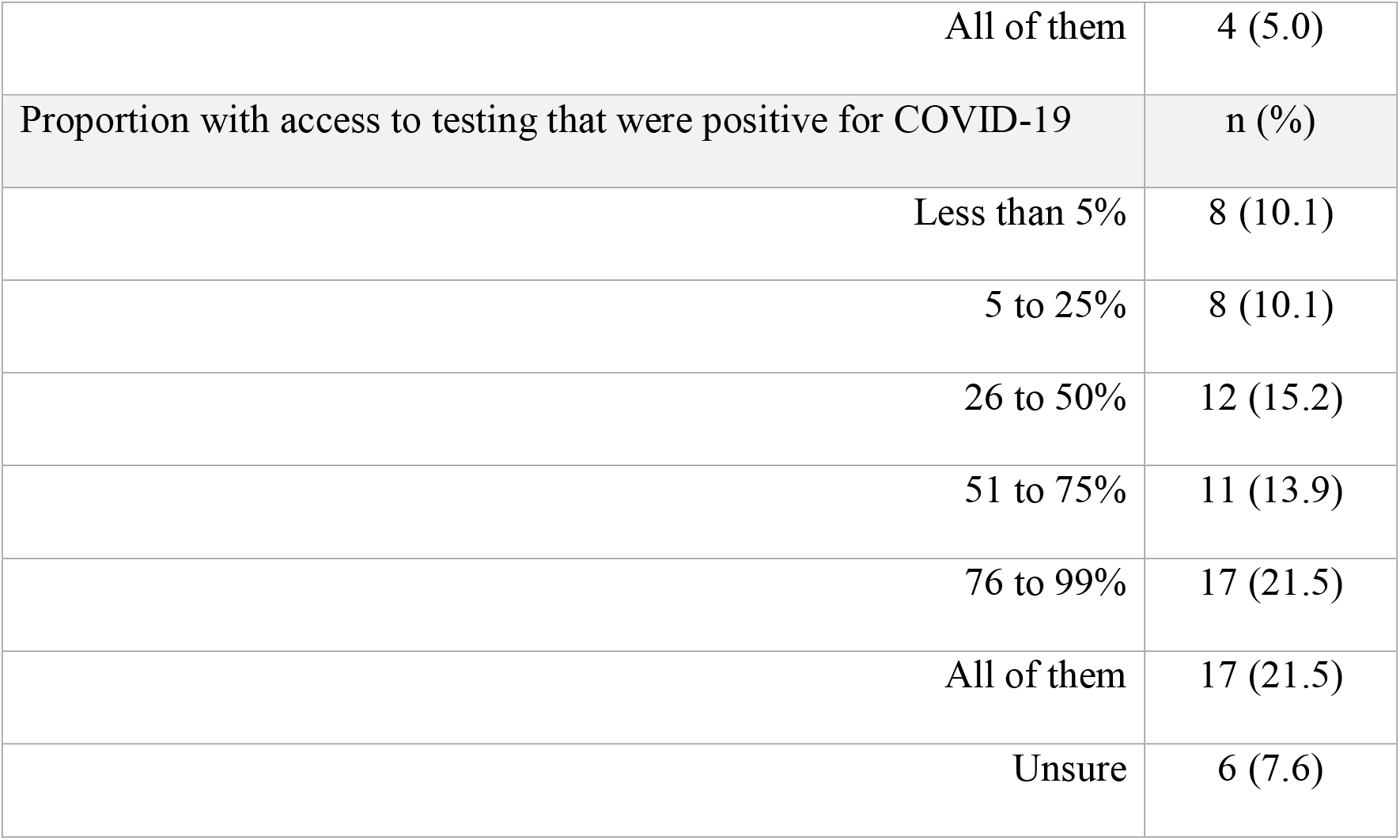
Number of patients treated for possible COVID-19 symptoms and COVID-19 testing status

| Approximate number of patients treated for possible COVID-19 symptoms | n (%) |
| --- | --- |
| Less than 10 | 29 (33.3) |
| 10 to 20 | 27 (33.8) |
| 21 to 30 | 10 (12.5) |
| 31 to 40 | 4 (5.0) |
| 41 to 50 | 2 (2.5) |
| More than 50 | 8 (10.0) |
| Proportion treated during acute initial phase of COVID-19 | n (%) |
| Less than 5% | 12 (15.0) |
| 5 to 25% | 15 (18.8) |
| 26 to 50% | 11 (13.8) |
| 51 to 75% | 10 (12.5) |
| 76 to 99% | 22 (27.5) |
| All of them | 10 (12.5) |
| Proportion starting treated without a positive test for COVID-19 | n (%) |
| Less than 5% | 33 (41.3) |
| 5 to 25% | 11 (13.8) |
| 26 to 50% | 18 (22.5) |
| 51 to 75% | 7 (8.8) |
| 76 to 99% | 7 (8.8) |
| All of them | 4 (5.0) |
| Proportion with access to testing that were positive for COVID-19 | n (%) |
| Less than 5% | 8 (10.1) |
| 5 to 25% | 8 (10.1) |
| 26 to 50% | 12 (15.2) |
| 51 to 75% | 11 (13.9) |
| 76 to 99% | 17 (21.5) |
| All of them | 17 (21.5) |
| Unsure | 6 (7.6) |

Table 4 presents data on how patient contact was undertaken. Most patients were seen by the participants either by phone (37%) or via videoconference (30%), with smaller numbers seen in-person, or contacted by email. Ten percent of the participants never closed their office, 20% closed and later reopened, and 6% made house calls. Seventeen percent of participants did in-person visits for post-acute COVID symptoms only. Seven percent of participants had difficulty obtaining personal protective equipment (PPE).

**Table 4.** How patient contact was undertaken and impact upon practice*

| Visit format for patients treated for possible COVID-19 symptoms | n (%) |
| --- | --- |
| In-person^ | 21 (17.1) |
| Phone | 45 (36.6) |
| Videoconference | 37 (30.1) |
| Email | 18 (14.6) |
| Combination | 30 (24.4) |
| Impact of COVID-19 on practice | n (%) |
| Never closed | 12 (9.8) |
| Closed and re-opened | 24 (19.5) |
| House calls for COVID symptoms | 7 (5.7) |
| In-person visits for acute COVID symptoms ONLY | 1 (0.8) |
| In-person visits for post-acute COVID symptoms ONLY | 21 (17.1) |
| In-person visits for any COVID symptoms | 12 (9.8) |
| Difficulty obtaining personal protective equipment (PPE) | 9 (7.3) |

\* Respondents could choose all that apply
^ In-person was in office, curbside, and outside

Table 5 indicates that the predominant form in which the CHM was taken by patients was granules (44%), followed by pill forms (33%). The duration of treatment was 30 or fewer days, with 11-20 days being the most common. Only two participants reported their patients experiencing side effects associated with the CHM, and this occurred in 5-25% of their patients treated with CHM for symptoms likely related to COVID-19. The severity of the side effects was rated as mild, and was associated with loose stools or constipation, nausea, and gagging. Forty-nine percent of participants experienced difficulty in obtaining the Chinese herbs that they needed for patient treatment during the COVID-19 pandemic.

**Table 5.** Form of Chinese herbs prescribed, duration of treatment, incidence of side effects, difficulty obtaining Chinese herbs

| Chinese herb formulations prescribed for possible COVID-19 symptoms* | n (%) |
| --- | --- |
| Raw herbs | 23 (18.7) |
| Granules | 52 (42.3) |
| Patent formulas (pill) | 40 (32.5) |
| Differed by patient | 5 (4.1) |
| Differed within patient | 15 (12.2) |
| Other^ | 8 (6.5) |
| Main Chinese herb formulation for COVID-19 symptoms | n (%) |
| Raw herbs | 11 (14.1) |
| Granules | 34 (43.6) |
| Patent formulas (pill) | 26 (33.3) |
| Other | 7 (9.0) |
| Duration of treatment | n (%) |
| Less than 10 days | 23 (29.5) |
| 11-20 days | 26 (33.3) |
| 21-30 days | 20 (25.6) |
| 31-60 days | 8 (10.3) |
| 61-120 days | 1 (1.3) |
| 121-180 days | 0 (0) |
| More than 6 months | 0 (0) |
| Side effects from Chinese herbs for COVID-19 symptoms | n (%) |
| Yes | 2 (2.6) |
| No | 76 (97.4) |
| Proportion with side effects | n (%) |
| Less than 5% | 0 |
| 5-25% | 2 |
| 26-50% | 0 |
| 51-75% | 0 |
| More than 75% | 0 |
| Severity of side effects were mainly<sup>Δ</sup> | n (%) |
| Severe | 0 |
| Moderate | 0 |
| Mild | 2 |
| Difficulty getting Chinese herbs | n (%) |
| Yes<sup>Σ</sup> | 38 (48.7) |
| No | 40 (51.3) |

\* Respondents could choose all that apply
<sup>Δ</sup> Other was pre-boiled vacuum packs, tinctures, and cooking recipes
<sup>Δ</sup> Side effects reported were loose stools/constipation, nausea, gagging
<sup>Σ</sup> Increased demand, supply chain and delivery issues

Table 6 presents data related to the sources of information and their use in devising CHM formulas for patients with symptoms likely related to COVID-19. Many different sources of information were consulted in addition to their own clinical experience that was used by 50% of the participants. However, only two participants used primarily their own clinical experience alone. High frequency sources were continuing education providers (31%), colleagues (31%), Chinese medicine journals (28%), herbal medicine companies (27%), and notes from continuing education courses (25%). Sixty percent of the participants used information about the use of CHM for past viral outbreaks (SARS, Zika, Ebola, H1N1). Participants predominantly (24% of participants) used a combination of herbal formulas that had been created pre- and post-COVID-19, which they had modified or not. Nineteen percent of participants mainly used pre-COVID non-modified formulas, and 18% of participants mainly used modified versions of formulas that were created to treat COVID-19 symptoms. Thirteen percent of providers used formulas that they designed themselves. The majority were influenced in their creation and prescribing of CHM formulas by the anecdotal information about the use of Chinese herbs to treat COVID-19 patients that came from China (69%), and by the scientific studies examining the use of CHM to treat COVID-19 (60%).

**Table 6.** Sources of information used and influences in devising Chinese herbal formulas

| Sources of information for devising Chinese herbal medicine formulas* | n (%) |
| --- | --- |
| Text books | 38 (30.9) |
| Notes from coursework | 17 (13.8) |
| Notes from continuing education courses | 31 (25.2) |
| Herbal medicine companies | 33 (26.8) |
| Professional publications | 19 (15.5) |
| Continuing education providers | 38 (30.9) |
| Biomedical journals | 19 (15.5) |
| Chinese medicine journals | 34 (27.6) |
| Colleagues | 38 (30.9) |
| Clinical experience | 62 (50.4) |
| Primarily clinical experience alone | 2 (1.6) |
| Other^ | 9 (7.3) |
| Use of information from past viral outbreaks to inform Chinese herbal medicine prescribing | n (%) |
| Yes<sup>Δ</sup> | 47 (60.3) |
| No | 31 (39.7) |
| Use and modification of preexisting Chinese herbal medicine prescriptions | n (%) |
| Mainly formulas that were not modified from sources created pre-COVID | 15 (19.2) |
| Mainly formulas that were not modified from sources describing the treatment of COVID infected patients | 6 (7.7) |
| Mainly formulas that were modified from sources created pre-COVID | 8 (10.3) |
| Mainly formulas that were modified from sources describing the treatment of COVID infected patients | 14 (18.0) |
| Mainly formulas that were designed by the provider | 10 (12.8) |
| Combination of above | 19 (24.4) |
| Other<sup>Σ</sup> | 6 (7.7) |
| Influenced by anecdotal information from China about use of herbs to treat COVID-19? | n (%) |
| Yes | 54 (69.2) |
| No<sup>Ω</sup> | 24 (30.8) |
| Influenced by scientific studies examining the use of herbs to treat COVID-19? | n (%) |
| Yes | 47 (60.3) |
| No | 31 (39.7) |

\*Respondents could choose all that apply
^Other included research from China, and the use of classical texts
Δ27/47 (56%) used it because there was no specific information about the use of Chinese herbs for COVID at the time
ΣUse of patent formulas and classical Chinese herbal medicine formulas
ΩFive responded in the open-ended section for this question that they had consulted with mentors, colleagues, or professors who had knowledge of the use of Chinese herbs for COVID-19 in China

Sixty-three percent of participants reported that the outcomes associated with CHM treatment of their patients was predominantly undertaken through discussions with them about their symptoms (Table 7). Ten participants used outcome instruments, and of these, four participants used them for all of their patients, and two participants used them for more than 75% of their patients.

**Table 7.**
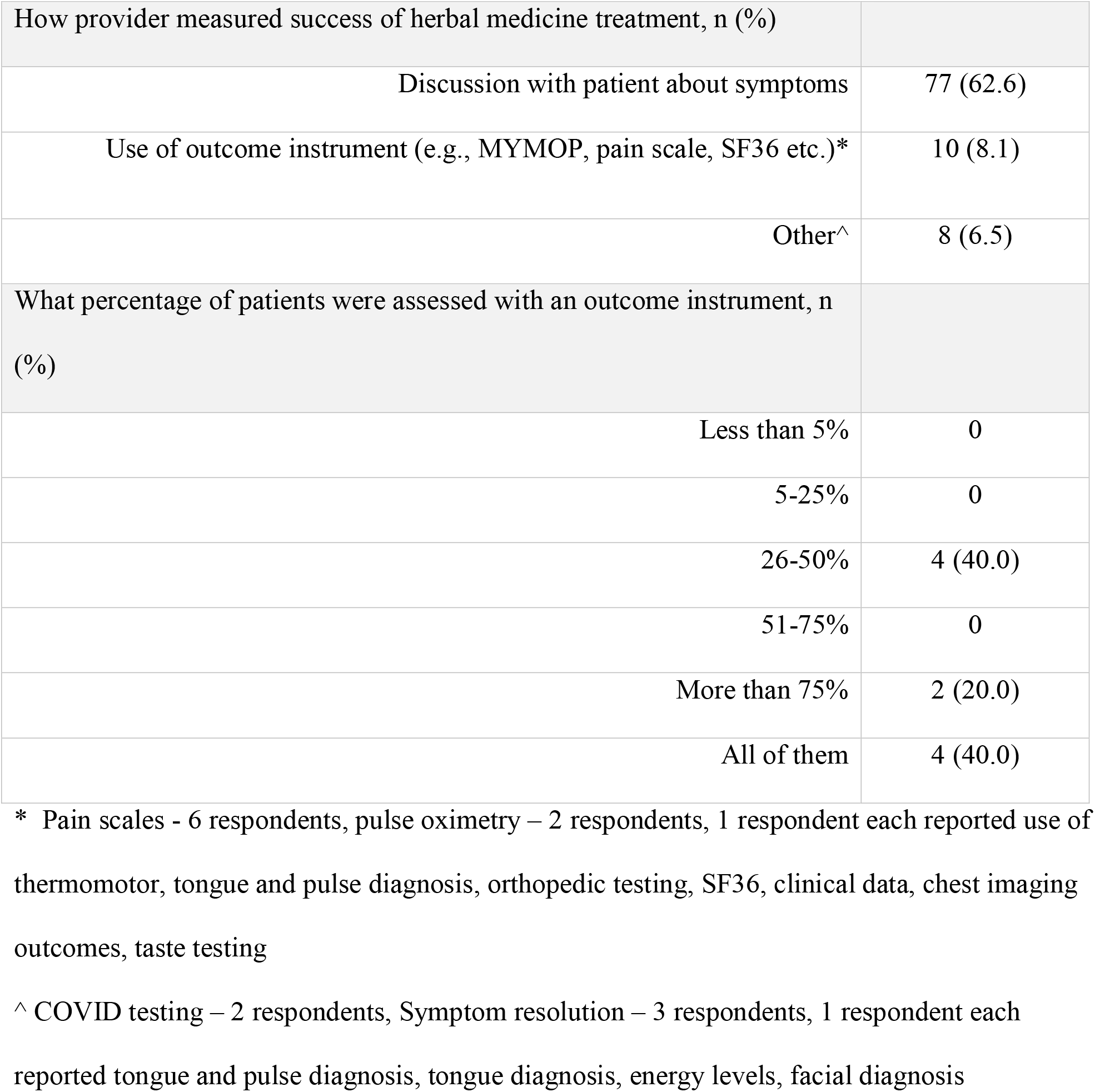
Measurement of treatment success

| How provider measured success of herbal medicine treatment, n (%) |  |
| --- | --- |
| Discussion with patient about symptoms | 77 (62.6) |
| Use of outcome instrument (e.g., MYMOP, pain scale, SF36 etc.)* | 10 (8.1) |
| Other^ | 8 (6.5) |
| What percentage of patients were assessed with an outcome instrument, n (%) |  |
| Less than 5% | 0 |
| 5-25% | 0 |
| 26-50% | 4 (40.0) |
| 51-75% | 0 |
| More than 75% | 2 (20.0) |
| All of them | 4 (40.0) |

\* Pain scales - 6 respondents, pulse oximetry – 2 respondents, 1 respondent each reported use of thermomotor, tongue and pulse diagnosis, orthopedic testing, SF36, clinical data, chest imaging outcomes, taste testing
^ COVID testing – 2 respondents, Symptom resolution – 3 respondents, 1 respondent each reported tongue and pulse diagnosis, tongue diagnosis, energy levels, facial diagnosis

Most of the patients that the participants were treating for symptoms likely related to COVID-19 were not also receiving biomedical treatments (Table 8). Ninety-seven percent of participants reported that they had no patients die of COVID-19. The majority of participants reported that 25% or less of their patients developed long hauler syndrome (post-acute sequelae SARS-CoV-2 infection - PASC).

**Table 8.**
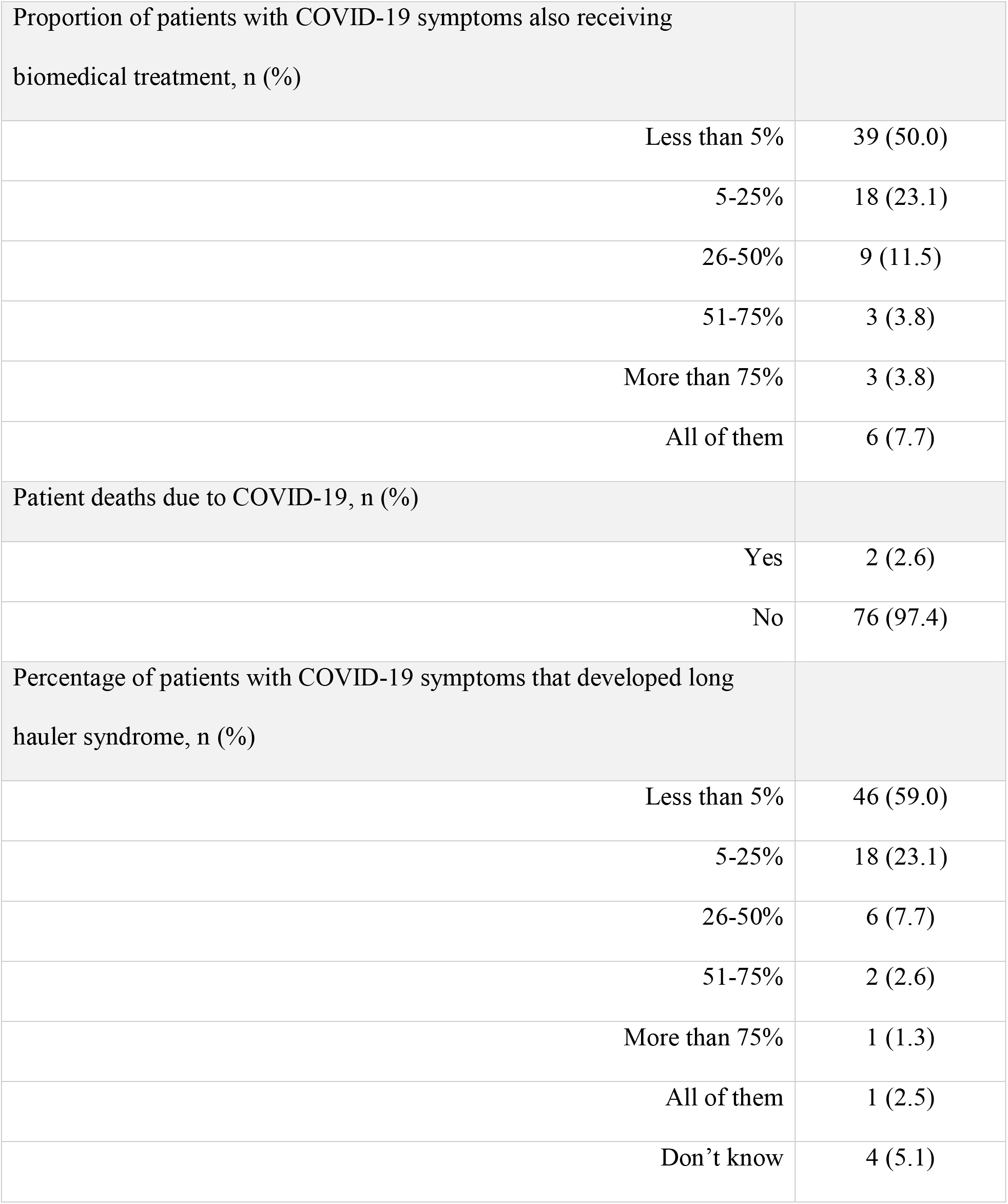
Proportion of patients also receiving biomedical treatments, who died, and who developed long-COVID

There were no significant associations between whether participants had received formal research or evidence-based medicine training and whether they were influenced by the scientific studies, used biomedical journals to inform their creation or choice of CHM formulas, or used outcome instruments to assess the success of the CHM medicine treatments (Table 9). We also found no association between participants’ years of clinical experience and whether they used their own clinical experience to devise CHM formulas, or between years of experience and whether they modified CHM formulas or designed their own formulas (Table 10).

**Table 9.**
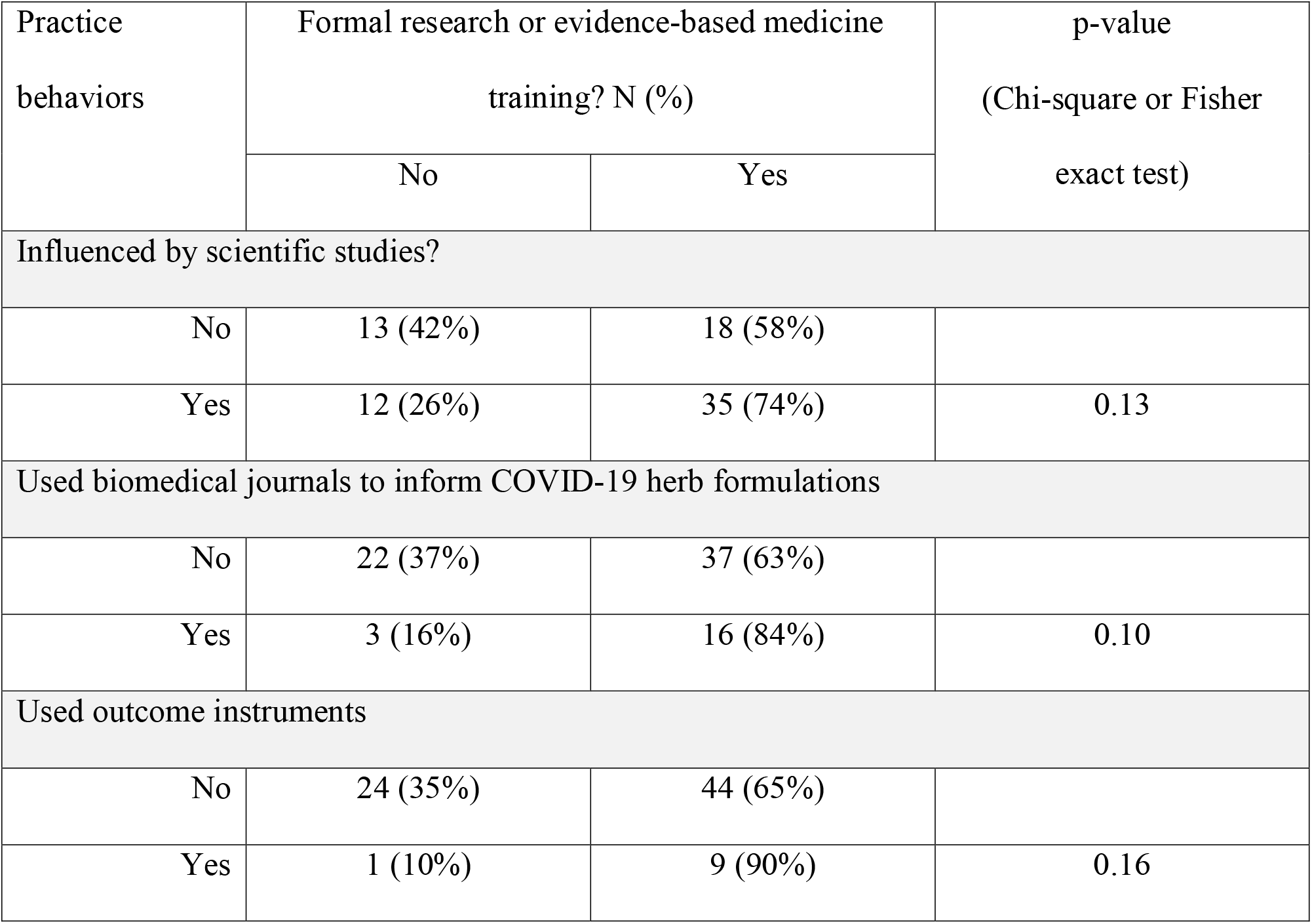
Associations between formal research training and COVID-19 practice behaviors

**Table 10.** Associations between years of clinical experience and CHM practice behaviors

| Practice behaviors | Years of clinical experience<sup>#</sup> |  | p-value<br>(t-test or ANOVA F-test) |
| --- | --- | --- | --- |
|  | N | Mean<br>(SD) |  |
| Used clinical experience for COVID-19 herb formulations |  |  | 0.12 |
| No | 38 | 14.8<br>(10.0) |  |
| Yes | 62 | 18.1<br>(10.3) |  |
| COVID-19 formulas (Q18) |  |  | 0.07 |
| Did not modify from other sources | 21 | 13.6 (7.1) |  |
| Modified from other sources | 22 | 19.5<br>(11.5) |  |
| Designed themselves | 10 | 20.3 (7.5) |  |

## Discussion

Our survey collected data from 103 licensed acupuncturists with on average 17 years of clinical experience who prescribed CHM to patients with symptoms likely related to COVID-19. Our participants came from all geographical regions across the US. About a third of participants may have been COVID-19 infected themselves, and of these 97% (all but one person) took CHM medicine for their symptoms. Contrary to popular belief^28^ the majority of participants (65%) had received the COVID-19 vaccine. This exceeded the proportion of the US population that had been vaccinated at the end of the study period – 57.2% for a single dose and 50.9% for two doses,^29^ and was similar to the proportion of hospital-based US healthcare workers that had been vaccinated by that time.^30^ Patients were treated in both the acute initial phase of COVID-19 infection and in later stages, including patients with long hauler syndrome (post-acute sequelae SARS-CoV-2 infection - PASC). Most patients were known to have COVID-19 by virtue of a positive COVID-19 test. Our study suggests that for the majority of the patients treated by the participants, CHM was the only treatment provided by a licensed healthcare practitioner to which these patients had access.

Seventeen percent of our participants saw patients with symptoms likely related to COVID-19 in person and 10% never closed their offices. This difference reflects the use of open-air locations including curbside treating patients in cars and outdoor clinics, as well as house calls. The most commonly used telehealth mechanism was the phone, which was similar to that reported in a survey of US acupuncturists investigating the use of telehealth during the COVID-19 pandemic.^24^ Their study also reported that of the 1,045 respondents, only 103 had prescribed CHM for patients likely infected with COVID-19. This suggests that our sample size, although small, may be representative of the licensed acupuncturists who were prescribing CHM during the pandemic.

Granulated and pill forms of CHM were the most common forms prescribed, as would be expected because these are the easiest and most convenient for patients to consume. As was reported in studies conducted in China,^23^ the CHM was usually taken for 20 days or less with little side effects. Almost half of the participants experienced difficulty obtaining the Chinese herbs and this was likely related to greatly increased demand due to usage in China and other Asian countries,^9,31,2^ and pandemic related interruptions to the supply chain, and delivery services being delayed.

Participants reported using a wide variety of different sources of information to inform their thinking around prescribing CHM for their patients with symptoms likely related to COVID-19. Such sources ranged from anecdotal information obtained from colleagues in the US and other countries, textbooks, continuing education sources, coursework notes, and from both Chinese medicine and biomedical journals. These sources cover all categories in the evidence pyramid^32^ with a predominance for the lower rungs of the pyramid – expert opinions, textbooks and observational research. Most of the scientific studies that had been published by the end of the time when the survey was undertaken were case studies.^33,23^ As such, at that time there was limited access to randomized controlled trials (RCTs) and systematic reviews.

This situation created a unique set of circumstances with regard to practicing evidence-based medicine. When evidence exists, sources highest on the evidence pyramid, RCTs and systematic reviews, are the superior sources. Given that the majority of the study participants (68%) had had training in research or evidence-based medicine, and that 60% of them were influenced by the scientific studies examining the use of CHM to treat COVID-19, we speculate that had high-quality RCTs and systematic reviews existed they would have been viewed as a reliable evidence source and been utilized.

Given the necessity to use evidence sources lower on the evidence pyramid, our data shows a concerted effort by the participants to access as broad a range of information sources as possible. The extensive information about the use of CHM for past viral epidemics including SARs, and H1N1 influenza^6,9^ was used by 60% of the participants. Fifty-six percent of these participants reported that they used this information because there was no specific information about the use of CHM for COVID-19 at the time.

There did not appear to be any association between those that had research and evidence-based medicine training and the use of scientific studies and biomedical journals. This may have been because of the lack of information from scientific sources relative to the large amount of information that was disseminated throughout the US about the use of CHM to treat COVID-19 infected patients in China.^19^ The latter was also directly applicable to clinical practice, unlike many research studies where internal validity is often more important than external validity. The lack of clinical applicability to real-world acupuncture practice has been reported as a deterrent to the use of scientific research by licensed acupuncturists in previous studies.^34,35,36^ The importance of this may have been magnified by the extreme and unusual circumstances associated with the early stages of the pandemic where no biomedical treatments were available.

Survey participants reported that the majority of their patients were not receiving biomedical treatment. Considering death was a possible outcome, applying the treatment that had the best supporting evidence at that time, which was CHM, would have likely been seen as the most ethical and best choice.

Half of the participants also used their own clinical experience; an essential component of evidence-based medicine practice, as originally defined by David Sackett.^37^ However, only two people exclusively used their own clinical experience. The use of clinical experience was also reflected in the majority of the participants either modifying existing CHM formulas or designing their own. This reflects a high level of confidence and experience with designing CHM formulas among the majority of the participants, as we might expect given that the average time in practice was 17 years.

The death rate of patients treated by the participants appeared very low with only two participants reporting that patients they had treated died. The development of long hauler syndrome (post-acute sequelae SARS-CoV-2 infection – PASC) was also low with 82% of participants reporting that less than 25% of their patients had developed long-hauler syndrome. The incidence of long-hauler syndrome among COVID-19 infected individuals is estimated to be between 33-60%.^38,39,40^ Our study detailing the use of CHM for symptoms likely related to COVID-19, along with the systematic reviews and metanalyses published after our study was completed^31,9^ suggesting that CHM may be an effective treatment for COVID-19 infection, supports the need for further investigation.

## Limitations

There were several limitations to this study. We do not have conclusive data on how many licensed acupuncturists were treating patients with symptoms likely related to COVID-19 with CHM during this stage of the pandemic, and so are unable to verify how representative our sample was. The survey used in this study was not psychometrically tested.

## Conclusions

Our study shows that licensed acupuncturists were prescribing CHM for patients who were infected with COVID-19, and for many such patients this was the only therapeutic intervention they had access to through a licensed healthcare provider. The majority of participants in our study had received the COVID-19 vaccine, accessed information about the use of CHM from journal articles and scientific studies as well as anecdotal sources, and assessed outcomes associated with their treatments in a systematic way. This reflects compliance with an evidence-based medicine approach to patient care and to protecting themselves against COVID-19 infection. Given the lack of biomedical treatments for COVID-19 it is possible that their efforts contributed to their patients’ recovery from COVID-19. Although at the time unproven through high-quality RCTs, CHM represented a possibly viable approach to treating COVID-19 infection according to the observational research that existed at that time. The participants in our study demonstrated a keen ability to make use of existing evidence to diligently contribute to a major public health crisis.

## Data Availability

All data produced in the present study are available upon reasonable request to the authors.

## Acknowledgments

We are grateful to the licensed acupuncturists who participated in our study and their dedicated efforts to continue to provide care for patients throughout the COVID-19 pandemic, which at times put their own health at risk. We also thank the many colleagues and professional organizations who assisted in the dissemination of our survey for participant recruitment.

## Contributions

All authors contributed to the design of the survey, data analysis, and the writing of this manuscript.

## Funding

This study was not supported by any grant funding.

## Competing Interests

None declared

## Patient consent for publication

Not required

## Ethics approval

This study was approved by the Institutional Review Board of Albert Einstein College of Medicine, IRB # 2020-12556. Consent was obtained from participants at initiation of the online survey.

## Provenance and per review

Not commissioned; externally peer reviewed

## Data availability statement

Data available on request

## Checklist/Reporting Statement

We used the STROBE cross sectional checklist when writing our report.^41^

## Patient and Public Involvement

It was not appropriate or possible to involve patients or the public in the design, or conduct, or reporting, or dissemination plans of our research.

## Notes

### Competing Interest Statement

The authors have declared no competing interest.

### Funding Statement

This study did not receive any funding.

### Author Declarations

The Institutional Review Board of Albert Einstein College of Medicine gave ethical approval for this work.

## References

1. Lee DYW, Li QY, Liu J, et al. Traditional Chinese herbal medicine at the forefront battle against COVID-19: Clinical experience and scientific basis. Phytomedicine 2021;80:153337. doi: 10.1016/j.phymed.2020.153337 [published Online First: 20200928]

2. Lyu M, Fan G, Xiao G, et al. Traditional Chinese medicine in COVID-19. Acta Pharm Sin B 2021;11(11):3337–63. doi: 10.1016/j.apsb.2021.09.008 [published Online First: 2021/09/20]

3. Sun CY, Sun YL, Li XM. The role of Chinese medicine in COVID-19 pneumonia: A systematic review and meta-analysis. Am J Emerg Med 2020;38(10):2153–59. doi: 10.1016/j.ajem.2020.06.069 [published Online First: 20200708]

4. Scheid V. Transmitting Chinese medicine: Changing perceptions of body, pathology, and treatment in late imperial China. Asian Med (Leiden) 2013;8(2):299–360.

5. Liu J, Manheimer E, Shi Y, et al. Chinese herbal medicine for severe acute respiratory syndrome: a systematic review and meta-analysis. J Altern Complement Med 2004;10(6):1041–51. doi: 10.1089/acm.2004.10.1041

6. Luo H, Tang QL, Shang YX, et al. Can Chinese Medicine Be Used for Prevention of Corona Virus Disease 2019 (COVID-19)? A Review of Historical Classics, Research Evidence and Current Prevention Programs. Chin J Integr Med 2020;26(4):243–50. doi: 10.1007/s11655-020-3192-6 [published Online First: 20200217]

7. Yang Y, Islam MS, Wang J, et al. Traditional Chinese Medicine in the Treatment of Patients Infected with 2019-New Coronavirus (SARS-CoV-2): A Review and Perspective. Int J Biol Sci 2020;16(10):1708–17. doi: 10.7150/ijbs.45538 [published Online First: 20200315]

8. Zhang B LS, Zhang J. Syndrome in TCM and therapeutic effect of integrated medicine in patients with SARS. Tianjin Journal of Traditional Chinese Medicine 2004;6

9. Zhang T, Li X, Chen Y, et al. Evidence Mapping of 23 Systematic Reviews of Traditional Chinese Medicine Combined With Western Medicine Approaches for COVID-19. Front Pharmacol 2021;12:807491. doi: 10.3389/fphar.2021.807491 [published Online First: 20220207]

10. Pang WT, Jin XY, Pang B, et al. [Analysis on pattern of prescriptions and syndromes of traditional Chinese medicine for prevention and treatment of COVID-19]. Zhongguo Zhong Yao Za Zhi 2020;45(6):1242–47. doi: 10.19540/j.cnki.cjcmm.20200218.502

11. Birch S, Lee MS, Alraek T, et al. Overview of Treatment Guidelines and Clinical Practical Guidelines That Recommend the Use of Acupuncture: A Bibliometric Analysis. J Altern Complement Med 2018;24(8):752–69. doi: 10.1089/acm.2018.0092 [published Online First: 20180618]

12. Miller DW, Roseen EJ, Stone JAM, et al. Incorporating Acupuncture Into American Healthcare: Initiating a Discussion on Implementation Science, the Status of the Field, and Stakeholder Considerations. Global Advances in Health and Medicine 2021;10:21649561211042574. doi: 10.1177/21649561211042574

13. Candon M, Nielsen A, Dusek JA. Trends in Insurance Coverage for Acupuncture, 2010-2019. JAMA Netw Open 2022;5(1):e2142509. doi: 10.1001/jamanetworkopen.2021.42509 [published Online First: 20220104]

14. Bensky D, Clavey S, Stöger E. Chinese Herbal Medicine: Materia Medica: Eastland Press 2004.

15. Unschuld PU. Medicine in China: A history of ideas.. 1985

16. Gao R-r, Hu Y-t, Dan Y, et al. Chinese herbal medicine resources: Where we stand. Chinese Herbal Medicines 2020;12(1):3–13. doi: https://doi.org/10.1016/j.chmed.2019.08.004

17. Scheid V, Bensky D, Barolet R. Chinese Herbal Medicine: Formulas & Strategies: Eastland Press 2009.

18. Chen JK, Chen TT. Chinese Herbal Formulas and Applications: Pharmacological Effects & Clinical Research: Art of Medicine Press 2009.

19. eLotus. TCM Resources for Coping with COVID-19 2021, September 21 [Available from: https://www.elotus.org/content/tcm-resources-covid-19.

20. eLotus. John Chen, Pharm.D., Ph.D., O.M.D., L.Ac. 2022, January 1 [Available from: https://www.elotus.org/bio/john-chen-pharmd-phd-omd-lac.

21. Chen JK, Chen TT, Crampton L. Chinese Medical Herbology and Pharmacology: Art of Medicine Press 2004.

22. National Certification Commission for Acupuncture and Oriental Medicine. National Certification Commission for Acupuncture and Oriental Medicine 2022, January 1 [Available from: https://www.nccaom.org/.

23. Xiong X, Wang P, Su K, et al. Chinese herbal medicine for coronavirus disease 2019: A systematic review and meta-analysis. Pharmacol Res 2020;160:105056. doi: 10.1016/j.phrs.2020.105056 [published Online First: 20200702]

24. Lee TL, Langley BO, Noborikawa J, et al. The Acupuncture and Telehealth Survey: A Cross-Sectional Survey Exploring Early COVID-19 Impacts on the Acupuncture Profession. J Integr Complement Med 2022;28(1):36–44. doi: 10.1089/jicm.2021.0151

25. Bolboacă SD, Jäntschi L, Sestraş AF, et al. Pearson-Fisher Chi-Square Statistic Revisited. Information 2011;2(3):528–45.

26. Stata Statistical Software: Release 13 [program]: StataCorp LLC, 2013.

27. United States Census Bureau. Census Regions and Divisions of the United States 2022, June 23 [Available from: https://www2.census.gov/geo/pdfs/maps-data/maps/reference/us_regdiv.pdf.

28. Coulter ID, Whitley MD, Khorsan R, et al. Incorporating Complementary and Integrative Health Providers in the Public Health Pandemic Response: Lessons from COVID-19 and Recommendations for the Future from a Multidisciplinary Expert Panel. Santa Monica, CA: RAND Corporation 2022.

29. Centers for Disease Control and Prevention. COVID Data Tracker 2022, June 23 [Available from: https://covid.cdc.gov/covid-data-tracker/#vaccination-trends.

30. Reses HE, Jones ES, Richardson DB, et al. COVID-19 vaccination coverage among hospital-based healthcare personnel reported through the Department of Health and Human Services Unified Hospital Data Surveillance System, United States, January 20, 2021-September 15, 2021. Am J Infect Control 2021;49(12):1554–57. doi: 10.1016/j.ajic.2021.10.008 [published Online First: 20211118]

31. An X, Zhang Y, Duan L, et al. The direct evidence and mechanism of traditional Chinese medicine treatment of COVID-19. Biomed Pharmacother 2021;137:111267–67. doi: 10.1016/j.biopha.2021.111267 [published Online First: 2021/01/14]

32. Burns PB, Rohrich RJ, Chung KC. The levels of evidence and their role in evidence-based medicine. Plast Reconstr Surg 2011;128(1):305–10. doi: 10.1097/PRS.0b013e318219c171

33. López-Alcalde J, Yan Y, Witt CM, et al. Current State of Research About Chinese Herbal Medicines (CHM) for the Treatment of Coronavirus Disease 2019 (COVID-19): A Scoping Review. J Altern Complement Med 2020;26(7):557–70. doi: 10.1089/acm.2020.0189 [published Online First: 20200624]

34. Ryan J. The use of evidence in acupuncture clinical practice. Australian journal of acupuncture and chinese medicine 2006;1(1):19–23.

35. Anderson BJ, Jurawanichkul S, Kligler BE, et al. Interdisciplinary Relationship Models for Complementary and Integrative Health: Perspectives of Chinese Medicine Practitioners in the United States. J Altern Complement Med 2019;25(3):288–95. doi: 10.1089/acm.2018.0268 [published Online First: 20181205]

36. Armour M, Betts D, Roberts K, et al. The Role of Research in Guiding Treatment for Women’s Health: A Qualitative Study of Traditional Chinese Medicine Acupuncturists. Int J Environ Res Public Health 2021;18(2) doi: 10.3390/ijerph18020834 [published Online First: 20210119]

37. Sackett DL, Rosenberg WM, Gray JA, et al. Evidence based medicine: what it is and what it isn’t. Bmj 1996;312(7023):71–2. doi: 10.1136/bmj.312.7023.71

38. Taquet M, Dercon Q, Luciano S, et al. Incidence, co-occurrence, and evolution of long-COVID features: A 6-month retrospective cohort study of 273,618 survivors of COVID-19. PLoS Med 2021;18(9):e1003773. doi: 10.1371/journal.pmed.1003773 [published Online First: 20210928]

39. Taquet M, Geddes JR, Husain M, et al. 6-month neurological and psychiatric outcomes in 236□379 survivors of COVID-19: a retrospective cohort study using electronic health records. Lancet Psychiatry 2021;8(5):416–27. doi: 10.1016/s2215-0366(21)00084-5 [published Online First: 20210406]

40. Huang C, Huang L, Wang Y, et al. 6-month consequences of COVID-19 in patients discharged from hospital: a cohort study. Lancet 2021;397(10270):220–32. doi: 10.1016/s0140-6736(20)32656-8 [published Online First: 20210108]

41. von Elm E, Altman DG, Egger M, et al. The Strengthening the Reporting of Observational Studies in Epidemiology (STROBE) statement: guidelines for reporting observational studies. J Clin Epidemiol 2008;61(4):344–9. doi: 10.1016/j.jclinepi.2007.11.008

